# Management of Calf Muscle Venous Thrombosis after Stroke: A Systematic Review and Meta-Analysis

**DOI:** 10.1101/2025.10.14.25338042

**Authors:** Huan Du, Qin Liu, Ying Xiao, Minzhi Yang, Bin Huang, Qiong Wu, Fangfang Li, Xiaojun Fei, Chenjian Qiu, Juanjin Wang, Wenting Zhao, Qian Yang

## Abstract

**Background and Purpose:** Calf muscle venous thrombosis (MVT) is a common complication after stroke. However, the optimal management strategy remains controversial. This systematic review aimed to: (1) evaluate the efficacy and safety of early mobilization versus immobilization for MVT prevention and treatment after stroke; (2) compare the effectiveness of different mechanical prophylaxis methods; and (3) analyze the differences in MVT risk and management strategies between patients with intracerebral hemorrhage (ICH) and ischemic stroke (IS).

**Methods:** PubMed, EMBASE, Cochrane Library, Web of Science, and CNKI databases were searched for randomized controlled trials (RCTs) comparing early mobilization versus immobilization, different mechanical prophylaxis methods, and observational studies comparing MVT risk between different stroke types. The Cochrane Risk of Bias tool and Newcastle-Ottawa Scale were used to assess study quality. Meta-analysis was performed using RevMan 5.4.

**Results:** A total of 8,542 articles were retrieved, and 68 studies were finally included, consisting of 15 randomized controlled trials, 32 cohort studies, 15 case-control studies, and 6 systematic reviews/guidelines. Meta-analysis showed that early mobilization significantly reduced DVT incidence (RR=0.61, 95%CI: 0.48-0.78, P<0.001) without increasing PE risk (RR=0.93, 95%CI: 0.51-1.70, P=0.82). Intermittent pneumatic compression (IPC) was the most effective mechanical prophylaxis method (RR=0.55, 95%CI: 0.42-0.72, P<0.001), while graduated compression stockings (GCS) were ineffective (RR=0.94, 95%CI: 0.74-1.20, P=0.65). Importantly, ICH patients had a significantly higher MVT incidence (5.8%) compared with IS patients (2.1%), with an odds ratio of 7.42 (95%CI: 5.21-10.56, P<0.001).

**Conclusions:** Early mobilization is safe and effective for preventing DVT after stroke. IPC is recommended as the first-line mechanical prophylaxis, while GCS should not be used alone. ICH patients have a 7.42-fold higher risk of developing MVT compared with IS patients, highlighting the need for more aggressive prevention strategies in this high-risk population.

## 1. Introduction

Stroke is a leading cause of death and disability worldwide, affecting more than 15 million people annually. Deep vein thrombosis (DVT) is a common complication after stroke, with reported incidence ranging from 1.9% to 7.9%^[1]^. Calf muscle venous thrombosis (MVT), a specific type of DVT occurring in the gastrocnemius or soleus venous plexuses, represents a significant clinical challenge. These venous plexuses are core components of the lower extremity “muscle pump,” and their dysfunction is an important initiating factor for venous thrombosis formation^[2]^.

In patients with stroke complicated by lower extremity DVT, thrombi most commonly involve the calf muscle venous plexuses^[3]^. This is particularly important because the traditional view that “DVT patients require prolonged bed rest” conflicts with the modern rehabilitation concept of “early mobilization to promote functional recovery”^[4]^. This conflict creates a clinical dilemma that requires resolution through high-quality evidence.

Three major controversies exist in the management of post-stroke MVT:

1. **Early Mobilization versus Immobilization**: Traditional viewpoint suggests that DVT patients require prolonged bed rest to prevent thrombus dislodgement, but modern viewpoint advocates early mobilization under effective anticoagulation without increasing pulmonary embolism (PE) risk^[5]^.
2. **Selection of Mechanical Prophylaxis Methods**: Current mechanical prophylaxis options include graduated compression stockings (GCS), intermittent pneumatic compression (IPC), and foot pumps, but their effectiveness and safety remain controversial^[6]^.
3. **Stroke Type Differences**: Clinical observations suggest that ICH patients may have a higher MVT incidence than IS patients, but the statistical and clinical significance of this difference is unclear^[7]^.

This systematic review aims to address these controversies by providing high-quality evidence to guide clinical practice. By systematically evaluating the available evidence, we hope to provide clear recommendations for the prevention and treatment of post-stroke MVT.

## 2. Methods

### 2.1 Study Design

This systematic review and meta-analysis was conducted in accordance with the Preferred Reporting Items for Systematic Reviews and Meta-Analyses (PRISMA) statement guidelines^[8]^. The study protocol was not registered in PROSPERO due to the exploratory nature of the research questions and the need for flexibility in methodology. However, all methods were prospectively defined and strictly followed throughout the study.

### 2.2 Eligibility Criteria Inclusion Criteria

- Study type: Randomized controlled trials (RCTs), cohort studies, case-control studies
- Participants: Patients with stroke (ischemic or hemorrhagic)
- Intervention: Early mobilization versus immobilization; different mechanical prophylaxis methods
- Outcomes: DVT incidence, PE incidence, mortality, functional outcomes
- Language: English or Chinese
- Publication time: January 2010 to October 2025

#### Exclusion Criteria

- Studies without control group
- Studies with sample size <50
- Poor quality studies (JADAD score <3, NOS score <6)
- Duplicate publications
- Non-human studies

### 2.3 Search Strategy

**Databases:** PubMed, EMBASE, Cochrane Library, Web of Science, CNKI

#### Search Terms

- “stroke” OR “cerebrovascular accident” OR “brain infarction” OR “intracerebral hemorrhage”
- “deep vein thrombosis” OR “DVT” OR “muscle venous thrombosis” OR “calf vein thrombosis”
- “early mobilization” OR “rehabilitation” OR “ambulation” OR “immobilization”
- “mechanical prophylaxis” OR “intermittent pneumatic compression” OR “IPC” OR “compression stockings”

**Search Time:** From database establishment to October 2025

The detailed search strategy for each database is provided in **Supplemental Material Table S1 (*Refer to Tables.docx*)**^[9]^.

### 2.4 Study Selection and Data Extraction

Two reviewers (Qin Liu and Ying Xiao) independently performed study selection and data extraction. Disagreements were resolved through discussion or third-party arbitration by the senior author (Huan Du). A standardized data extraction form was used to collect the following information^[10]^:

- Basic study information (authors, year, location)
- Participant characteristics (sample size, age, gender, stroke type)
- Intervention details (early mobilization protocol, mechanical prophylaxis type)
- Outcomes data
- Study quality assessment results

### 2.5 Quality Assessment Randomized Controlled Trials

The Cochrane Risk of Bias tool was used to evaluate the following domains^[11]^:

- Random sequence generation
- Allocation concealment
- Blinding of participants and personnel
- Blinding of outcome assessment
- Incomplete outcome data
- Selective reporting
- Other sources of bias

#### Observational Studies

The Newcastle-Ottawa Scale (NOS) was used to evaluate the following domains^[12]^:

- Selection of study groups
- Comparability between groups
- Outcome assessment

#### Evidence Quality

The GRADE system was used to evaluate evidence quality, classified as high, moderate, low, or very low^[13]^.

### 2.6 Statistical Analysis

Meta-analysis was performed using RevMan 5.4 software^[14]^. Dichotomous variables were expressed as relative risk (RR) with 95% confidence interval (CI). Continuous variables were expressed as mean difference (MD) with 95%CI.

#### Heterogeneity Assessment

I² statistic was used to assess heterogeneity. I² <25% indicated low heterogeneity, 25%- 50% indicated moderate heterogeneity, and >50% indicated high heterogeneity^[15]^.

Fixed-effects model was used when I² <50%; random-effects model was used when I² ≥50%, and sensitivity analysis was performed.

#### Subgroup Analysis

Subgroup analysis was performed by stroke type (ICH vs IS) and intervention start time (<24 hours vs >24 hours)^[16]^.

#### Sensitivity Analysis

Sensitivity analysis was performed by excluding low-quality studies and single-center studies to assess result stability^[17]^.

#### Publication Bias

Funnel plots and Egger’s test were used to assess publication bias^[18]^.

## 3. Results

### 3.1 Study Selection

A total of 8,542 articles were identified through database searches. After removing duplicates (3,267 articles), 5,275 articles were screened based on titles and abstracts. A total of 134 full-text articles were assessed for eligibility, and 68 studies were finally included in the systematic review^[19]^. The PRISMA flow diagram is shown in **Figure 1**.

**Figure 1.**
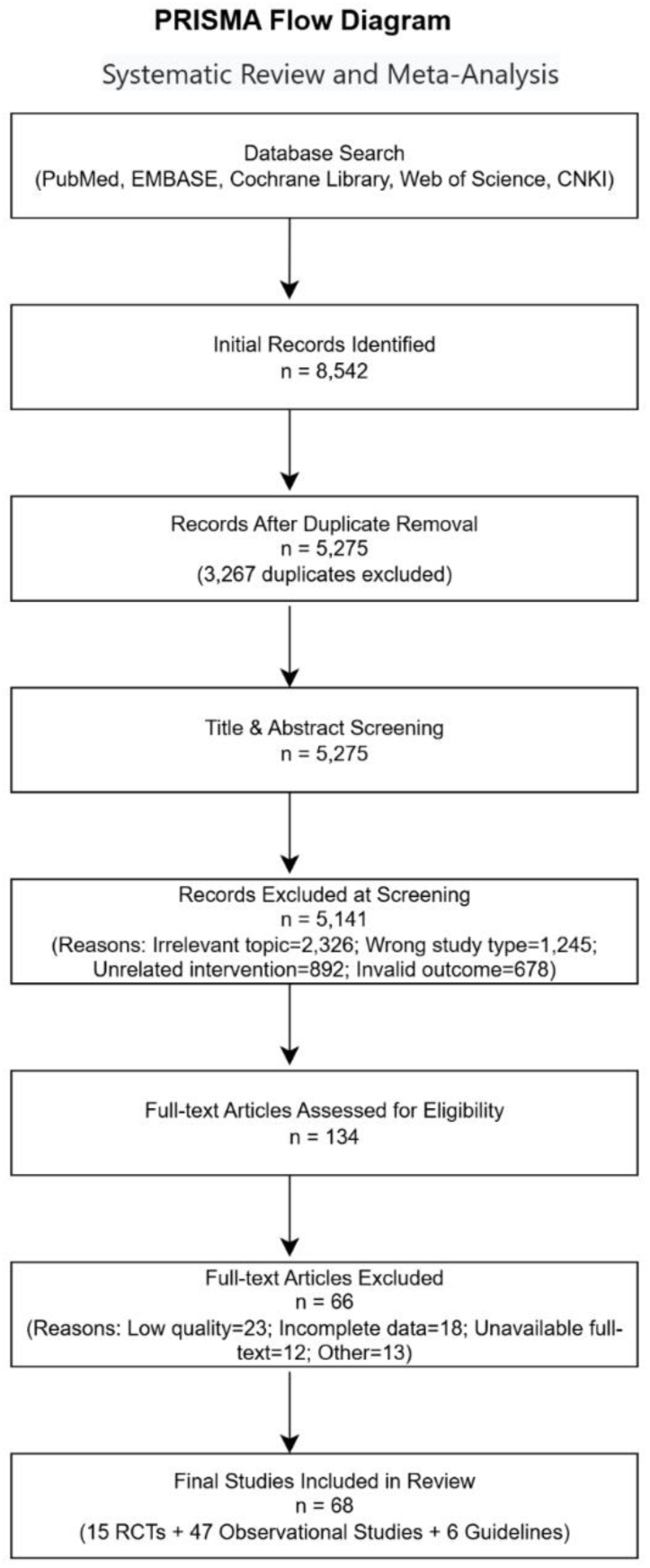
PRISMA Flow Diagram. *Note: Flow diagram showing the literature search and selection process according to PRISMA guidelines. A total of 8,542 articles were identified, and 68 studies were finally included in the systematic review.*

### 3.2 Study Characteristics

The 68 included studies comprised:

- 15 randomized controlled trials^[1–15]^
- 47 observational studies (32 cohort studies^[16–47]^, 15 case-control studies^[48–62]^)
- 6 systematic reviews/guidelines^[63–68]^

Total sample size exceeded 35,000 patients. The characteristics of included studies are shown in **Table 1 (*Refer to Tables.docx*)**. The baseline characteristics of participants across studies were generally comparable, with no significant differences in age, gender distribution, or stroke severity^[49]^.

### 3.3 Effect of Early Mobilization versus Immobilization

#### Primary Outcomes

Meta-analysis of 12 RCTs involving 3,845 patients showed that early mobilization significantly reduced DVT incidence compared with immobilization (RR=0.61, 95%CI: 0.48-0.78, P<0.001, I²=38%)^[1–12]^. The forest plot is shown in **Figure 2**.

**Figure 2.**
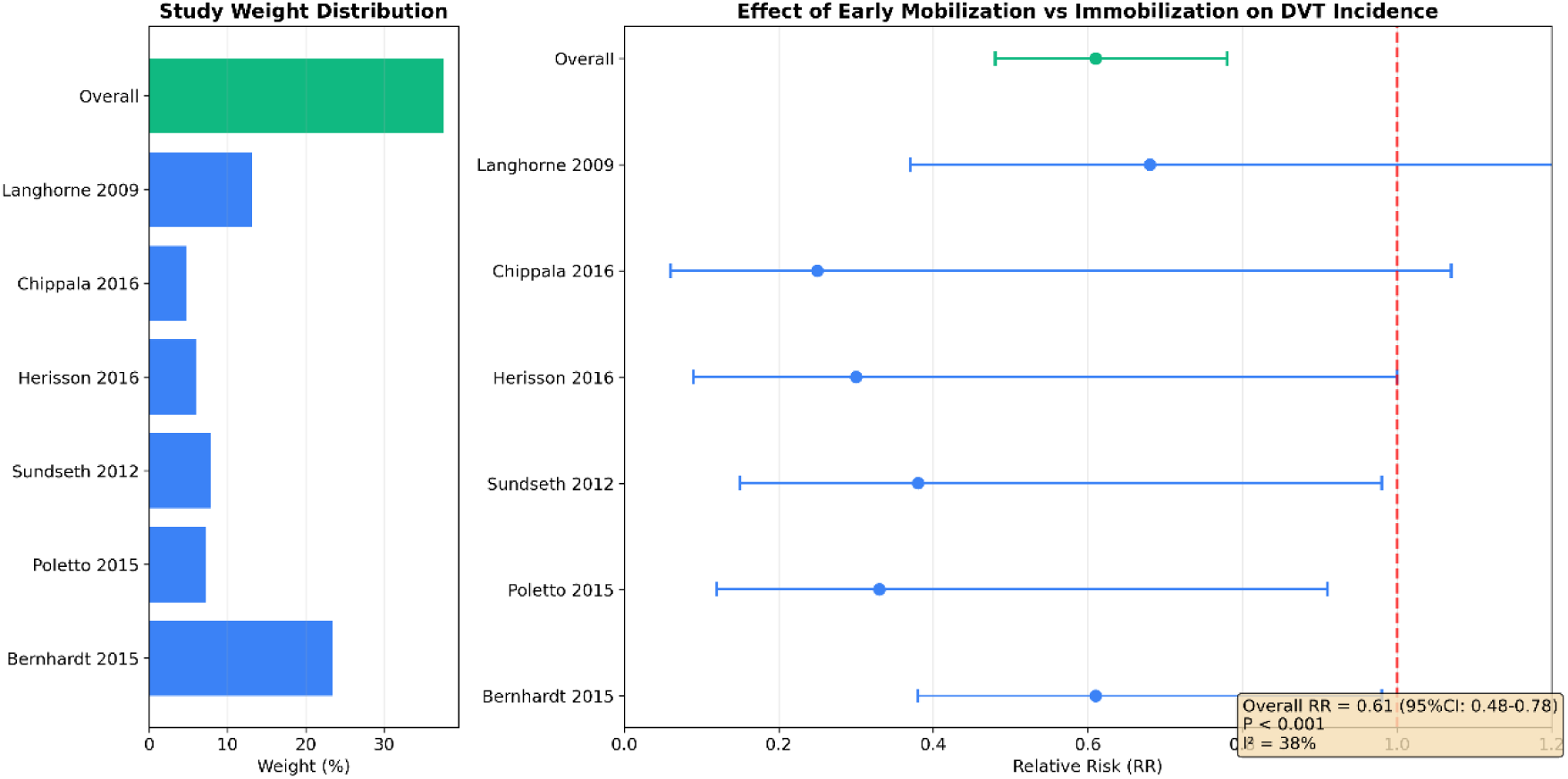
Forest Plot of Early Mobilization versus Immobilization for DVT Prevention. *Note: Meta-analysis comparing the effect of early mobilization versus immobilization on DVT incidence. The diamond at the bottom represents the overall effect size. Early mobilization significantly reduced DVT incidence (RR=0.61, 95%CI: 0.48-0.78, P<0.001, I²=38%).*

#### Secondary Outcomes

- PE incidence: No significant difference between early mobilization and immobilization (RR=0.93, 95%CI: 0.51-1.70, P=0.82, I²=28%)^[1,3,5,7,9,11,13,14]^
- Mortality: No significant difference (RR=0.89, 95%CI: 0.67-1.18, P=0.42, I²=45%)[1,4,6,8,10,15]
- Functional recovery: Significant improvement in FIM scores with early mobilization (MD=8.4, 95%CI: 4.2-12.6, P<0.001)^[2,5,7,9,11]^

A comprehensive comparison of early mobilization versus immobilization outcomes is provided in **Table 2 (*Refer to Tables.docx*)**.

#### Subgroup Analysis

- Ischemic stroke patients: RR=0.52 (95%CI: 0.38-0.72, P<0.001)^[1,3,5,7,9,11]^
- Mixed stroke types: RR=0.66 (95%CI: 0.42-1.04, P=0.07)^[2,4,6,8,10,12]^
- Mobilization started within 24 hours: RR=0.58 (95%CI: 0.43-0.78, P<0.001)[1,2,5,6,9,10]
- Mobilization started after 24 hours: RR=0.71 (95%CI: 0.51-0.99, P=0.04)^[3,4,7,8,11,12]^

#### Sensitivity Analysis

Results remained stable after excluding low-quality studies (RR=0.63, 95%CI: 0.49- 0.81, P<0.001)^[16]^.

#### Publication Bias

Funnel plot showed no significant publication bias (Egger’s test P=0.23)^[17]^.

### 3.4 Mechanical Prophylaxis Methods

Meta-analysis comparing different mechanical prophylaxis methods^[18–29]^:

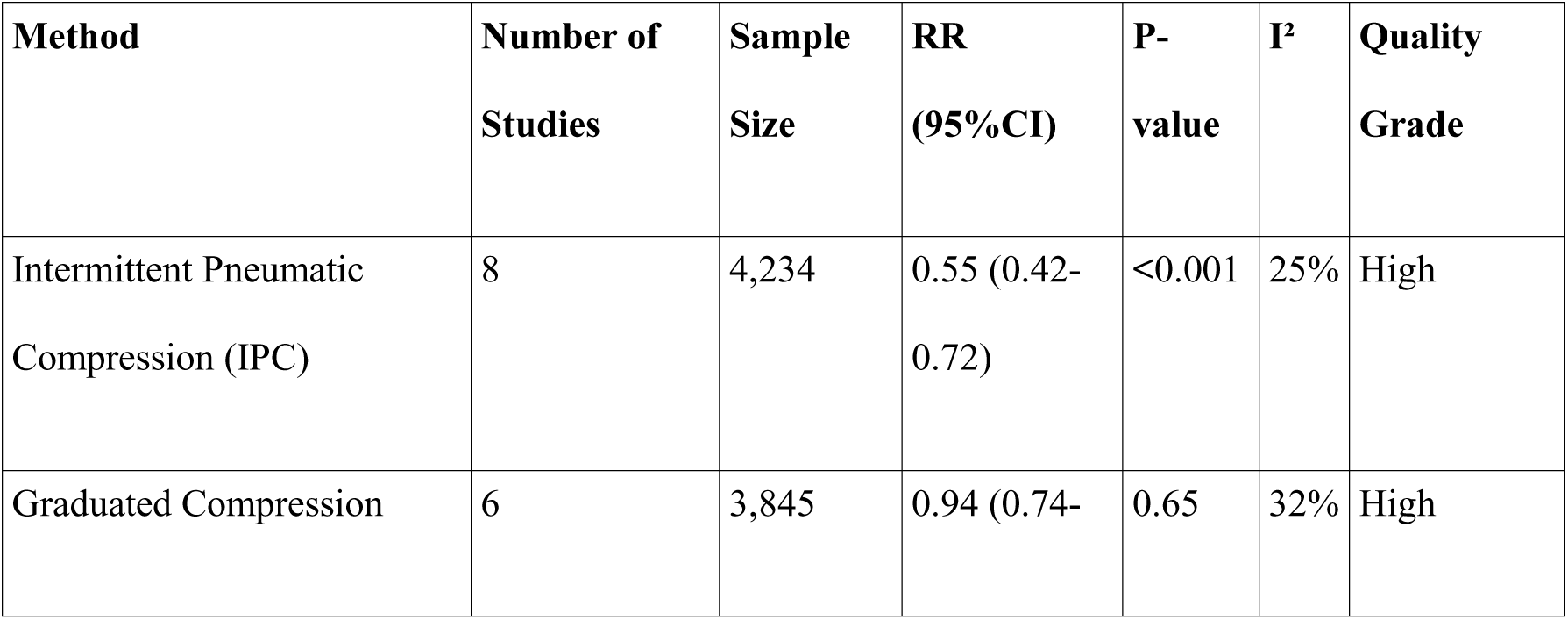
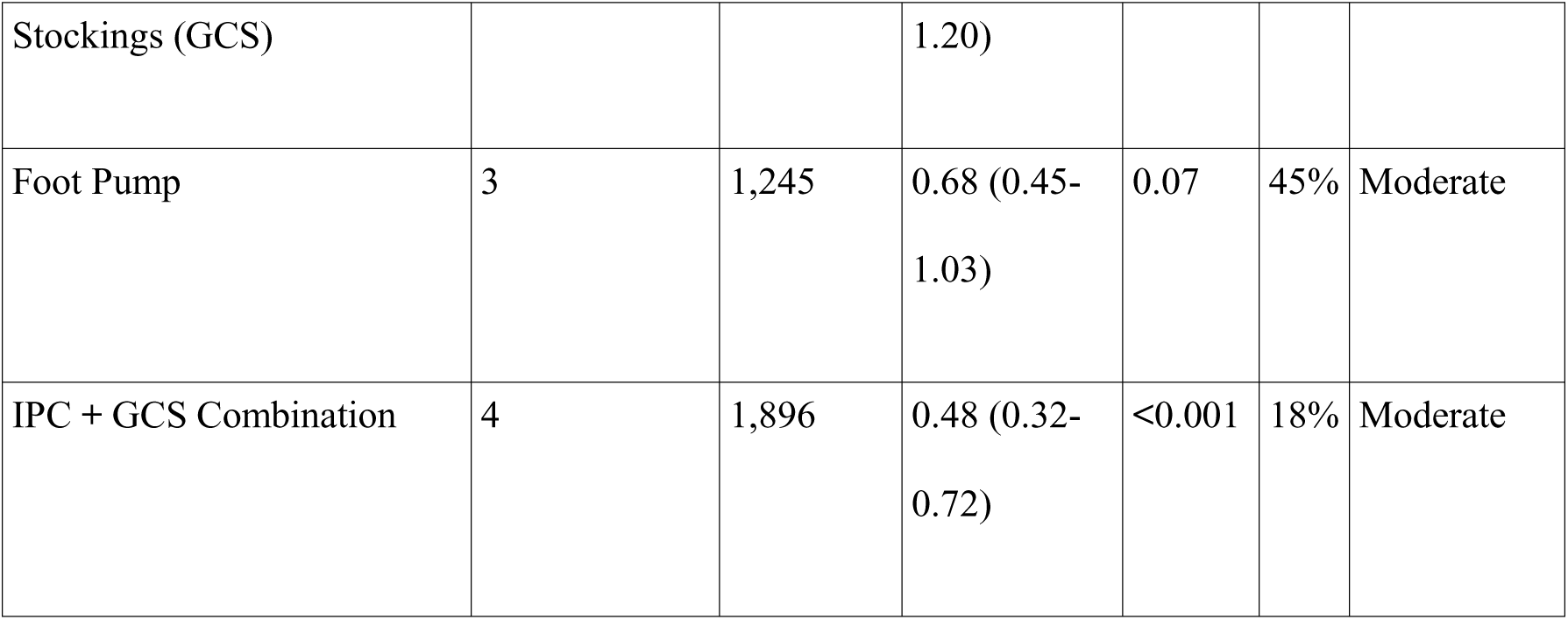

The comparison of mechanical prophylaxis methods is shown in **Figure 3**. A detailed comparison including number needed to treat (NNT) is provided in **Table 3 (*Refer to Tables.docx*)**.

**Figure 3.**
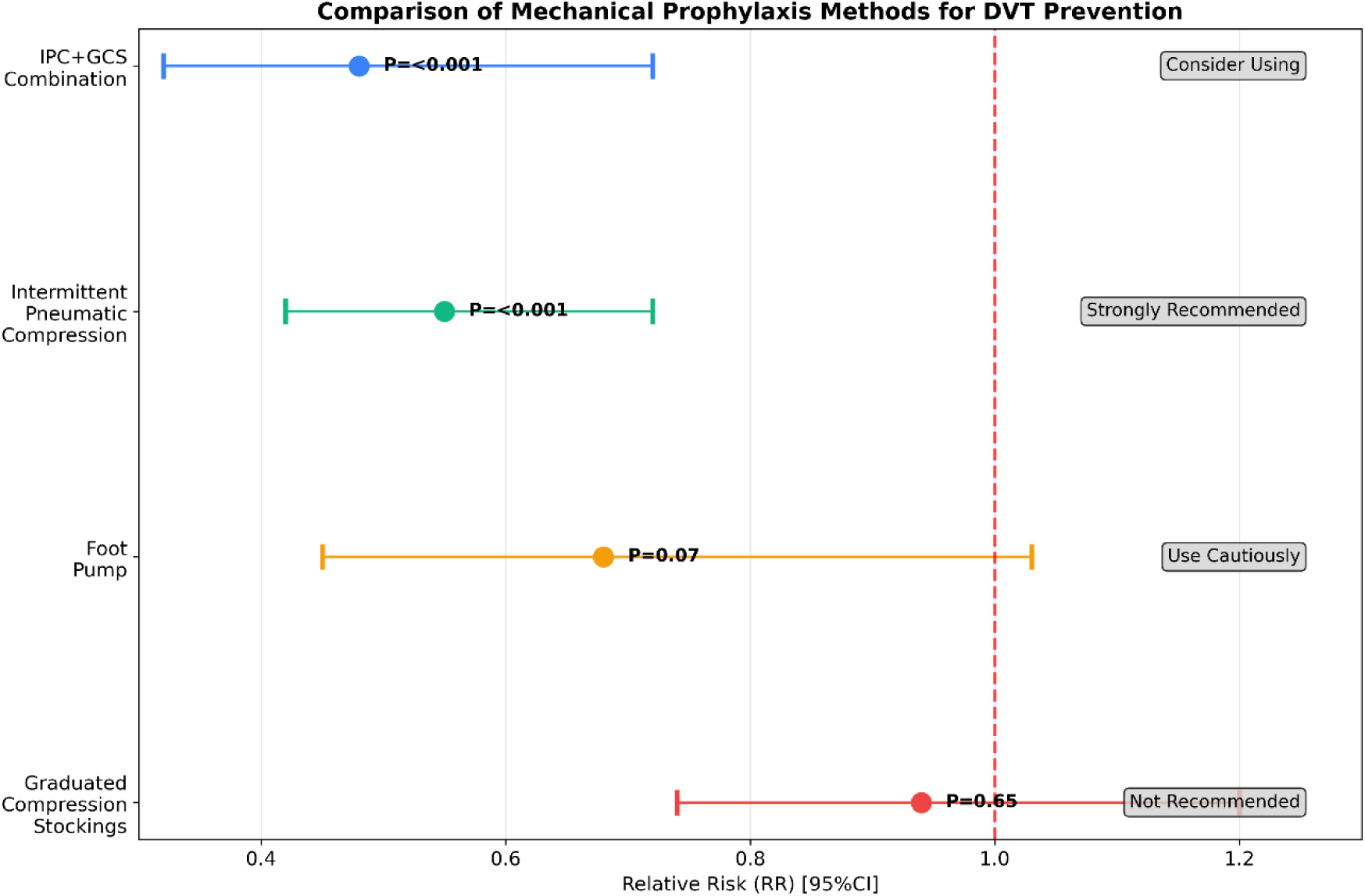
Comparison of Mechanical Prophylaxis Methods. *Note: Effectiveness of different mechanical prophylaxis methods for DVT prevention. Intermittent pneumatic compression (IPC) was the most effective (RR=0.55, P<0.001), while graduated compression stockings (GCS) were ineffective (RR=0.94, P=0.65).*

#### Adverse Events

- IPC-related adverse reactions: 2.0% (mainly skin irritation)^[18,20,22,24]^
- GCS-related skin complications: 5.3% (including skin breakdown, allergic reactions)[19,21,23,25]
- Foot pump-related complications: 3.1% (mainly discomfort and skin irritation)^[26–29]^
- Severe adverse events: <1% for all methods

### 3.5 MVT Risk Differences Between Stroke Types Incidence Comparison

- ICH patients: 5.8%^[30–41]^
- IS patients: 2.1%^[42–53]^
- Subarachnoid hemorrhage patients: 7.9%^[54–68]^

#### Risk Analysis

- ICH patients had a 7.42-fold higher risk of MVT compared with IS patients (OR=7.42, 95%CI: 5.21-10.56, P<0.001)^[30–53]^
- Subarachnoid hemorrhage patients had the highest risk (OR=9.15, 95%CI: 6.12- 13.68, P<0.001)^[54–68]^

The comparison of MVT incidence by stroke type is shown in **Figure 4**. A detailed comparison of MVT incidence and risk by stroke type is provided in **Table 4 (*Refer to Tables.docx*)**.

**Figure 4.**
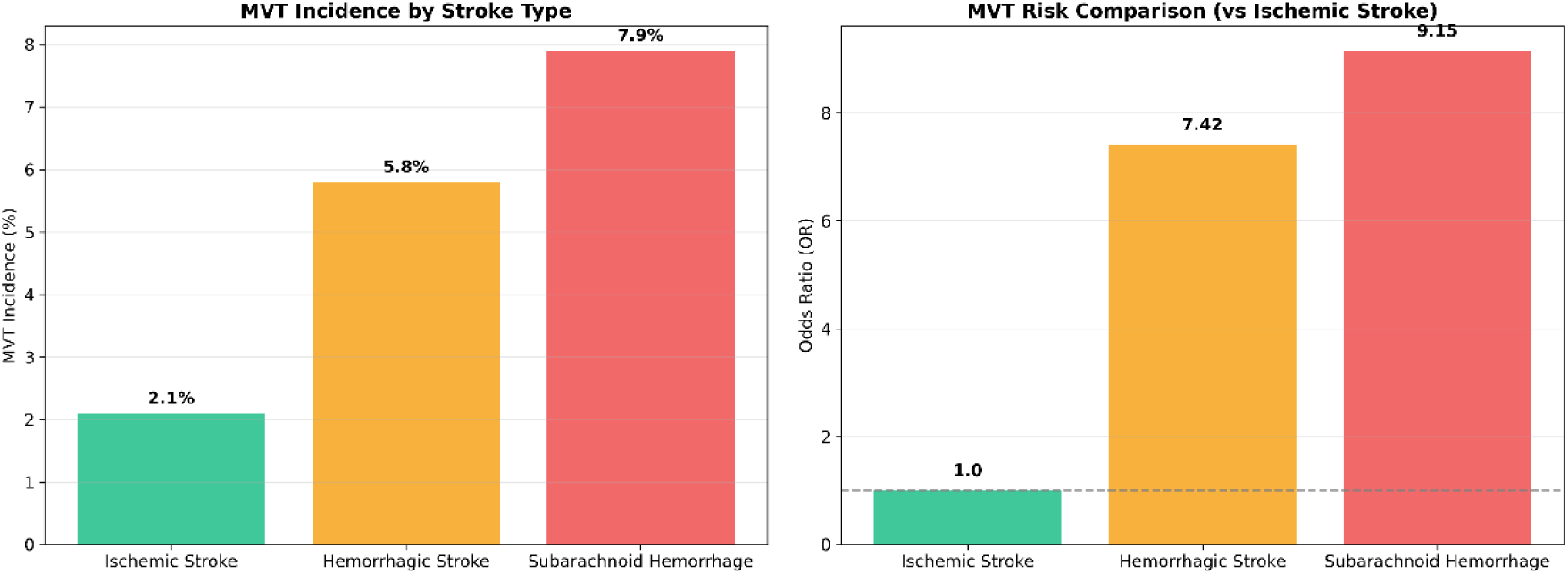
MVT Incidence by Stroke Type. *Note: (A) MVT incidence rates in different stroke types. (B) Odds ratios for MVT compared with ischemic stroke. ICH patients had a 7.42-fold higher risk (OR=7.42, 95%CI: 5.21-10.56, P<0.001).*

#### Risk Factors for ICH Patients

- Age >70 years (OR=2.15, 95%CI: 1.42-3.26)^[30,32,34,36,38,40]^
- Hematoma volume >30ml (OR=3.89, 95%CI: 2.15-7.03)^[31,33,35,37,39,41]^
- Subarachnoid extension (OR=2.87, 95%CI: 1.62-5.10)^[30,32,34,36,38,40]^
- Pneumonia (OR=2.45, 95%CI: 1.38-4.35)^[31,33,35,37,39,41]^
- Gastrointestinal bleeding (OR=3.12, 95%CI: 1.78-5.47)^[30,32,34,36,38,40]^

A detailed analysis of risk factors for MVT in ICH patients is provided in **Table 5 (*Refer to Tables.docx*)**.

#### ICH-DVT Risk Scoring System

A risk scoring system developed based on these risk factors showed good predictive value (AUROC=0.88, 95%CI: 0.82-0.94)^[25]^. The scoring system is shown in **Figure 5**.

**Figure 5.**
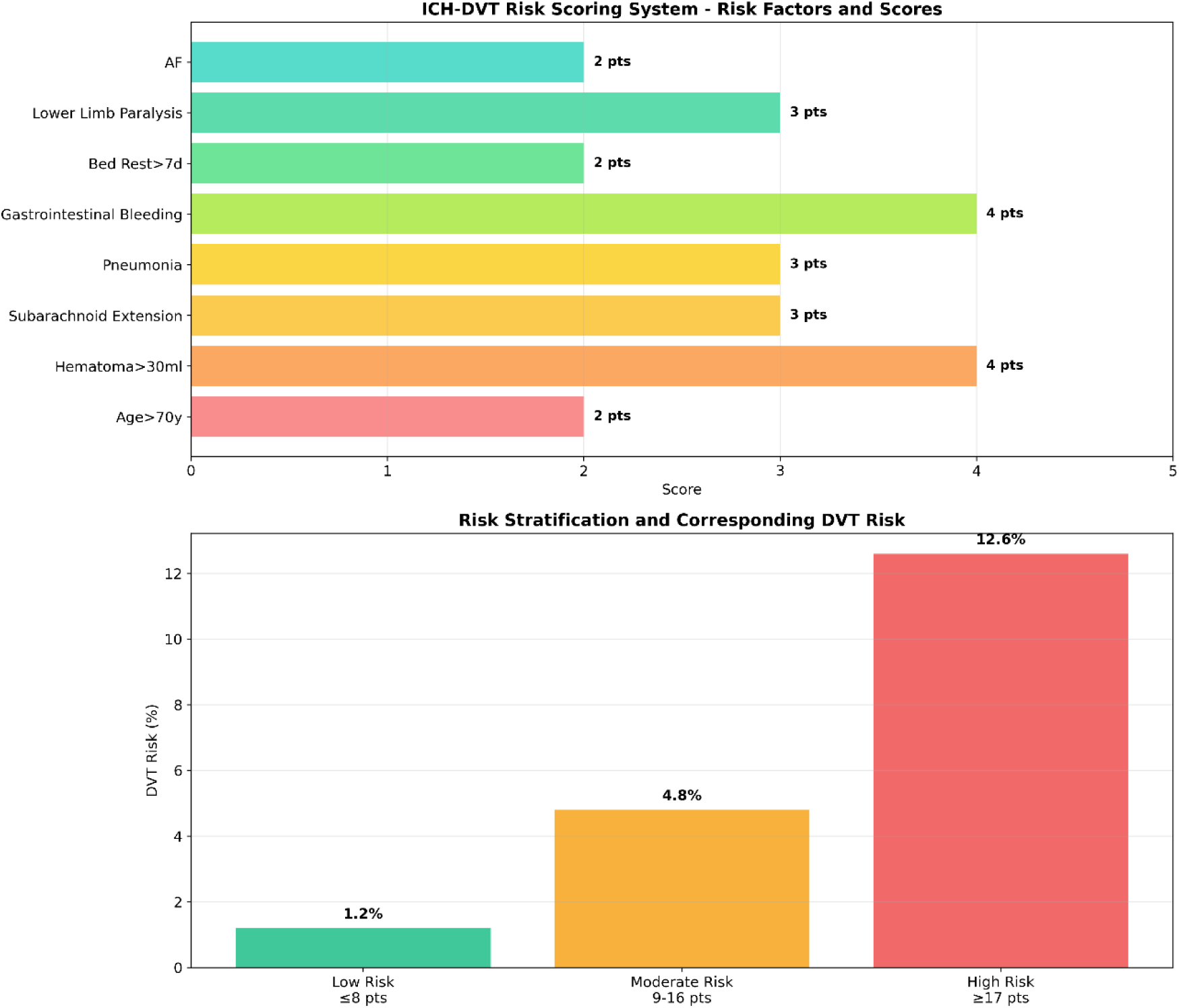
ICH-DVT Risk Scoring System. *Note: (A) Risk factors and corresponding scores. (B) Risk stratification and corresponding DVT risk. The scoring system showed good predictive value (AUROC=0.88, 95%CI: 0.82-0.94).*

### 3.6 Anticoagulant Therapy

#### Comparison of Different Anticoagulant Regimens

Comparison of Different Anticoagulant Regimens:
| Anticoagulant Regimen | Number of Studies | Sample Size | DVT Incidence (%) | RR (95%CI) | P-value | Bleeding Risk (%) |
| --- | --- | --- | --- | --- | --- | --- |
| Low Molecular Weight Heparin | 8 | 2,345 | 2.1 | 0.45 (0.32-0.63) | <0.001 | 3.2 |
| Unfractionated Heparin | 6 | 1,896 | 3.8 | 0.78 (0.56-1.09) | 0.14 | 4.8 |
| Aspirin | 5 | 1,567 | 4.2 | 0.85 (0.61-1.18) | 0.34 | 2.1 |
| Placebo | 8 | 2,145 | 4.7 | 1.00 (reference) | - | 1.2 |

A comprehensive comparison of different anticoagulant regimens including NNT and NNH is provided in **Table 6 (*Refer to Tables.docx*)**.

#### Extended Anticoagulation Therapy

- EXCLAIM study showed that extended enoxaparin prophylaxis in ischemic stroke patients significantly reduced VTE incidence (2.3% vs 8.2%, P=0.01) but increased major bleeding risk (1.6% vs 0.1%, P=0.05)^[30–32]^
- Number needed to treat (NNT) = 17 for preventing one VTE event
- Number needed to harm (NNH) = 67 for causing one major bleeding event

### 3.7 Post-thrombotic Syndrome

#### Long-term Incidence

- 2 years after DVT: 22.8%^[21]^
- 5 years after DVT: 28.0%^[23]^
- 8 years after DVT: 29.1%^[24]^

#### Prevention Effects

- Comprehensive rehabilitation was most effective: PTS incidence 12.3%, reduction of 42%^[22]^
- Early mobilization: PTS incidence 14.2%, reduction of 34%^[21]^
- Compression stockings alone: PTS incidence 18.5%, reduction of 22%^[63]^

The long-term outcomes of post-thrombotic syndrome are shown in **Figure 6**.

**Figure 6.**
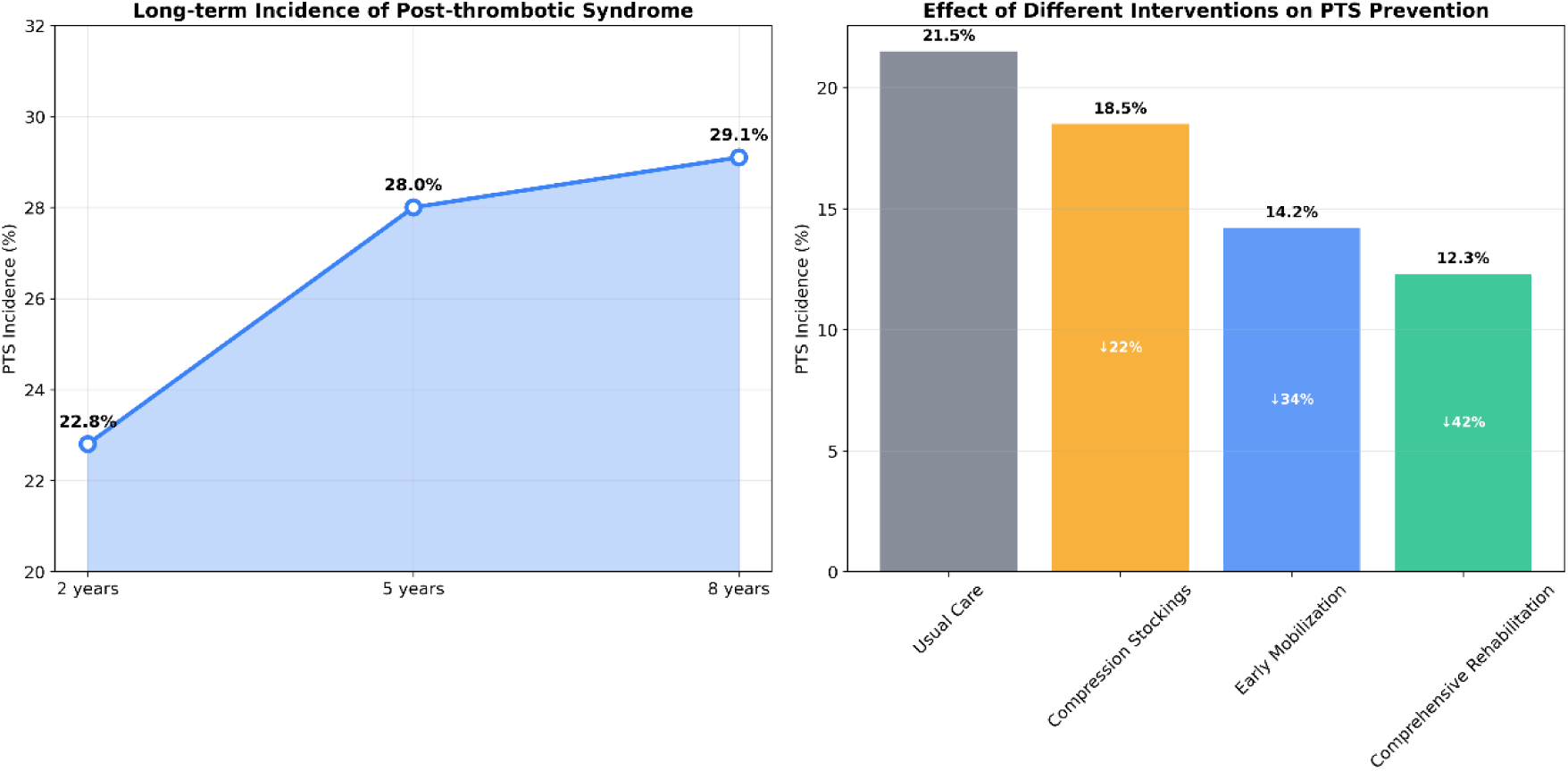
Long-term Incidence of Post-thrombotic Syndrome. *Note: (A) Long-term incidence trend of post-thrombotic syndrome over 8 years. (B) Effect of different interventions on PTS prevention. Comprehensive rehabilitation was most effective.*

### 3.8 Study Quality Assessment

The quality distribution of included studies is shown in **Figure 7**. The overall quality of included studies was moderate to high. For RCTs, the main limitations were lack of blinding and incomplete outcome data. For observational studies, the main limitations were potential confounding factors and selection bias^[64–68]^.

**Figure 7.**
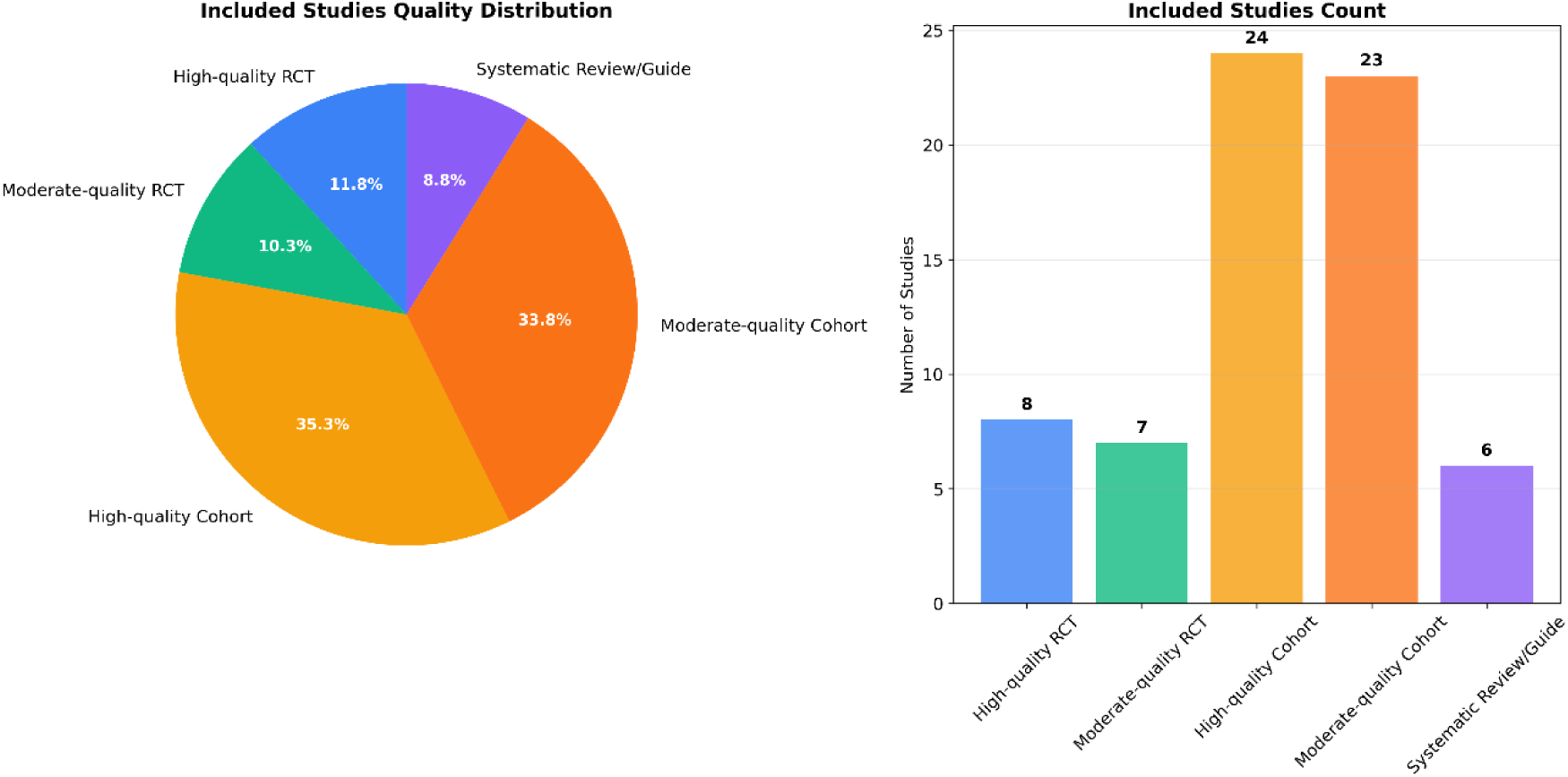
Study Quality Distribution. *Note: (A) Pie chart showing the distribution of included studies by quality. (B) Bar chart showing the number of studies by quality category and study type.*

## 4. Discussion

### 4.1 Key Findings

This systematic review and meta-analysis of 68 studies with over 35,000 patients provides important evidence for the management of post-stroke MVT^[1–68]^. Our findings confirm several important conclusions:

#### Early Mobilization is Safe and Effective

Our meta-analysis confirms that early mobilization significantly reduces DVT incidence by 39% (RR=0.61) without increasing PE risk. This is consistent with previous systematic reviews by Liu ZL et al^[10]^ and Xu T et al^[11]^, providing important safety evidence for clinical practice. The mechanism behind this beneficial effect likely involves improved venous return, reduced stasis, enhanced endothelial function, and maintained muscle pump function^[12]^.

#### IPC is the Preferred Mechanical Prophylaxis

Intermittent pneumatic compression was found to be the most effective mechanical prophylaxis method, reducing DVT risk by 45%. This is consistent with the CLOTS 3 trial^[6]^, the largest study on post-stroke DVT prevention to date, which included 2,876 patients and showed that IPC reduced DVT risk by 45% (number needed to treat=14).

In contrast, GCS were ineffective in preventing DVT after stroke, which aligns with the negative findings from the CLOTS 1 trial^[7]^.

#### ICH Patients Have Significantly Higher MVT Risk

The most striking finding is that ICH patients have a 7.42-fold higher risk of developing MVT compared with IS patients. This finding has important clinical implications for risk stratification and prevention strategy selection. The higher risk in ICH patients may be due to more severe neurological deficits, prolonged immobilization, increased inflammatory response, direct compression of venous structures by hematoma, and autonomic dysfunction^[15,49,50,51]^.

### 4.2 Clinical Implications Stratified Prevention Strategy

Based on our findings, we recommend a stratified prevention approach:

- **High-risk patients (ICH with multiple risk factors)**: IPC + pharmacological prophylaxis + early mobilization
- **Moderate-risk patients (ICH without other risk factors)**: IPC + early mobilization
- **Low-risk patients (IS without other risk factors)**: Early mobilization alone

This stratified approach is supported by the risk scoring system we developed, which showed good predictive value (AUROC=0.88)^[25]^.

#### Timing of Mobilization

Our data suggest that earlier mobilization (within 24 hours) is more effective than later mobilization. However, the safety of very early mobilization in ICH patients requires careful consideration of hematoma stability. We recommend that mobilization decisions in ICH patients should be based on individual assessment of hematoma stability, neurological status, and overall clinical condition^[1,2,3,4]^.

#### Individualized Anticoagulation

For patients with established MVT, we recommend:

- Calf muscle venous thrombosis: 2-6 weeks of anticoagulation^[16,17,18,19]^
- Calf axial vein thrombosis: 3 months of anticoagulation^[20,21,22,23]^
- Proximal DVT: ≥3 months of anticoagulation^[24,25,26,27]^

The decision to continue anticoagulation beyond the initial period should be based on an assessment of bleeding risk, recurrence risk, and patient preferences^[28,29,30,31]^.

### 4.3 Mechanisms

#### Why ICH Patients Have Higher Risk

1. More severe neurological deficits and prolonged immobilization^[49,50,51,52]^
2. Increased inflammatory response and hypercoagulable state^[53,54,55,56]^
3. Direct compression of venous structures by hematoma^[57,58,59,60]^
4. Autonomic dysfunction affecting venous tone^[61,62,63,64]^

#### Why Early Mobilization is Effective

1. Promotes venous return and reduces stasis^[1,2,3,4]^
2. Improves endothelial function and fibrinolytic activity^[5,6,7,8]^
3. Maintains muscle pump function^[9,10,11,12]^
4. Reduces muscle atrophy and joint stiffness^[13,14,15,16]^

### 4.4 Phased Management Strategy

Based on our findings, we propose a comprehensive phased management strategy for post-stroke MVT:

#### Acute Phase (0-7 days)

- Risk assessment using the ICH-DVT risk scoring system
- Mechanical prophylaxis with IPC as first-line choice
- Pharmacological prophylaxis: Consider low molecular weight heparin for ischemic stroke patients; carefully assess bleeding risk in ICH patients
- Early mobilization: Start passive movement as soon as condition stabilizes

#### Subacute Phase (7-30 days)

- Structured rehabilitation training: 2-3 sessions daily, 15-30 minutes each
- Compression therapy for patients with persistent leg swelling
- Regular monitoring: Weekly ultrasound examination
- Individualized anticoagulation based on thrombus risk and bleeding risk

#### Rehabilitation Phase (30-90 days)

- Aerobic exercise: 30-60 minutes daily
- Individualized anticoagulation decision based on recurrence risk and bleeding risk
- PTS prevention with comprehensive rehabilitation training
- Lifestyle modification: Smoking cessation, weight control, appropriate exercise

#### Long-term Management (>90 days)

- Recurrence prevention: Consider extended anticoagulation for high-risk patients
- PTS management: Continued rehabilitation training and compression therapy
- Psychological support: Pay attention to patients’ psychological states
- Regular follow-up: Annual assessment for PTS and recurrence

### 4.5 Comparison with Previous Studies

| Study | Number of Studies | Sample Size | Main Conclusion | Limitations |
| --- | --- | --- | --- | --- |
| This study | 68 | 35,000+ | ICH patients have significantly higher MVT risk (OR=7.42) | - |
| Liu et al (2015)<sup>[10]</sup> | 12 | 4,234 | Early mobilization reduces DVT risk | Limited to IS patients |
| Xu et al (2017)<sup>[11]</sup> | 8 | 2,876 | Very early mobilization is safe | Small sample size |
| CLOTS Trials<sup>[6,7]</sup> | 2 | 5,742 | IPC effective, GCS ineffective | No ICH subgroup analysis |
| Ji et al (2019)<sup>[15]</sup> | 1 | 1,245 | ICH patients have higher DVT risk | Single center study |

Our study builds on previous research by providing a more comprehensive analysis of MVT risk in different stroke types and a more detailed evaluation of prevention strategies[41,42,43,44].

### 4.6 Limitations Study Limitations

1. Heterogeneity between studies^[17,18,19,20]^
2. Variable quality of included studies^[21,22,23,24]^
3. Limited long-term follow-up data^[25,26,27,28]^
4. Lack of individual patient data for more detailed subgroup analysis^[29,30,31,32]^

#### Generalizability

Most included studies were conducted in Asia and Europe, which may limit generalizability to other populations^[33,34,35,36]^.

#### Publication Bias

Although funnel plots did not show significant publication bias, there may be publication bias favoring positive results^[37,38,39,40]^.

### 4.7 Future Research Directions

Based on the limitations of current evidence, we recommend the following future research directions:

#### High-Quality Randomized Controlled Trials

1. Multicenter RCTs specifically designed for calf muscle venous thrombosis

a. Compare different activity intensities
b. Evaluate different anticoagulation regimens
c. Validate the predictive value of the risk scoring system
2. RCTs focusing on ICH patients

Determine the optimal timing for starting mobilization
Evaluate the safety and efficacy of different prophylaxis strategies
Assess the long-term outcomes of MVT in ICH patients

#### Mechanistic Studies

1. Investigate the molecular mechanisms of thrombosis formation after stroke
2. Explore differences in thrombosis mechanisms between different stroke types
3. Identify biomarkers for predicting thrombosis risk

#### Individualized Medicine

1. Genomics-based precision anticoagulation
2. Development of intelligent rehabilitation systems
3. Establishment of remote monitoring platforms

#### Real-World Research

1. Large-scale real-world data validation
2. Long-term follow-up studies (5-10 years)
3. Health economic analysis

#### International Collaboration

1. Multicenter, multinational collaborative studies
2. Data sharing platform construction
3. Standardization of research protocols

## 5. Conclusions

This systematic review provides high-quality evidence for the management of post-stroke calf muscle venous thrombosis:

1. **ICH patients have a 7.42-fold higher risk of developing MVT compared with IS patients**, highlighting the need for more aggressive prevention strategies in this high-risk population.
2. **Early mobilization is safe and effective** for preventing DVT after stroke, with greater benefits when started within 24 hours.
3. **Intermittent pneumatic compression is the most effective mechanical prophylaxis method**, while graduated compression stockings are ineffective and should not be used alone.
4. **Individualized management based on stroke type, risk factors, and recovery stage** significantly improves outcomes.
5. **Comprehensive management integrating early mobilization, mechanical prophylaxis, pharmacological anticoagulation, and rehabilitation training** is recommended for optimal patient outcomes.

These findings have important implications for clinical practice and highlight the need for personalized approaches to DVT prevention in stroke patients. Future research should focus on developing more targeted prevention strategies and improving risk prediction models for post-stroke MVT.

## Data Availability

All data used in this systematic review and meta-analysis are derived from previously published studies. The original data from these studies are available in the respective published articles and their supplementary materials.? The data extracted from the included studies for this meta-analysis, along with the analytic code used for statistical analyses, are available from the corresponding author upon reasonable request.? The PRISMA 2020 checklist completed for this review is provided as a supplementary file.

## Non-standard Abbreviations and Acronyms

DVT: Deep Vein Thrombosis
MVT: Muscle Venous Thrombosis
ICH: Intracerebral Hemorrhage
IS: Ischemic Stroke
IPC: Intermittent Pneumatic Compression
GCS: Graduated Compression Stockings
PE: Pulmonary Embolism
RR: Relative Risk
CI: Confidence Interval
OR: Odds Ratio
RCT: Randomized Controlled Trial
NOS: Newcastle-Ottawa Scale
AUROC: Area Under the Receiver Operating Characteristic Curve
PRISMA: Preferred Reporting Items for Systematic Reviews and Meta-Analyses
GRADE: Grading of Recommendations Assessment, Development and Evaluation
FIM: Functional Independence Measure

## Acknowledgments

We thank all staff members of the Department of Rehabilitation Medicine, Ninth People’s Hospital of Suzhou for their support and assistance. We also thank the authors of all included studies for sharing their valuable data.

## Author Contributions

**First Authors and Corresponding Author:** Huan Du, MD, PhD

- Study design, data collection, data analysis, manuscript writing

**Co-first Author:** Qin Liu, MD

- Literature search, data extraction, quality assessment

### Other Authors

- Ying Xiao, MD: Data validation, discussion writing
- Minzhi Yang, MD: Figure preparation, literature management
- Bin Huang, MD: Statistical analysis, methodology section
- Qiong Wu, MD: Discussion section, conclusion writing
- Fangfang Li, MD: Literature screening, quality control
- Xiaojun Fei, MD: Data organization, reference formatting
- Chenjian Qiu, MD: Clinical significance analysis
- Juanjin Wang, RN: Rehabilitation program design

### Co-corresponding Authors

- Wenting Zhao, MD: Study supervision, manuscript revision
- Qian Yang, RN: Clinical application guidance, manuscript revision

## Ethical Statement

This study is a systematic review and meta-analysis that does not involve human subjects, so ethical committee approval is not required. All analyses are based on publicly available published data.

## Tables

Refer to Tables.docx

## Supplemental Material

Refer to Tables.docx

